# Artificial Intelligence-Based Analytics for Diagnosis of Small Bowel Enteropathies and Black Box Feature Detection

**DOI:** 10.1101/2020.08.06.20159152

**Authors:** Sana Syed, Lubaina Ehsan, Aman Shrivastava, Saurav Sengupta, Marium Khan, Kamran Kowsari, Shan Guleria, Rasoul Sali, Karan Kant, Sung-Jun Kang, Kamran Sadiq, Najeeha T. Iqbal, Lin Cheng, Christopher A. Moskaluk, Paul Kelly, Beatrice C. Amadi, S. Asad Ali, Sean R. Moore, Donald E. Brown

## Abstract

**Objectives:** Striking histopathological overlap between distinct but related conditions poses a significant disease diagnostic challenge. There is a major clinical need to develop computational methods enabling clinicians to translate heterogeneous biomedical images into accurate and quantitative diagnostics. This need is particularly salient with small bowel enteropathies; Environmental Enteropathy (EE) and Celiac Disease (CD). We built upon our preliminary analysis by developing an artificial intelligence (AI)-based image analysis platform utilizing deep learning convolutional neural networks (CNNs) for these enteropathies.

**Methods:** Data for secondary analysis was obtained from three primary studies at different sites. The image analysis platform for EE and CD was developed using convolutional neural networks (CNNs: ResNet and custom Shallow CNN). Gradient-weighted Class Activation Mappings (Grad-CAMs) were used to visualize the models’ decision making process. A team of medical experts simultaneously reviewed the stain color normalized images done for bias reduction and Grad-CAM visualizations to confirm structural preservation and biological relevance, respectively.

**Results:** 461 high-resolution biopsy images from 150 children were acquired. Median age (interquartile range) was 37·5 (19·0 to 121·5) months with a roughly equal sex distribution; 77 males (51·3%). ResNet50 and Shallow CNN demonstrated 98% and 96% case-detection accuracy, respectively, which increased to 98·3% with an ensemble. Grad-CAMs demonstrated models’ ability to learn distinct microscopic morphological features.

**Conclusion:** Our AI-based image analysis platform demonstrated high classification accuracy for small bowel enteropathies which was capable of identifying biologically relevant microscopic features, emulating human pathologist decision making process, performing in the case of suboptimal computational environment, and being modified for improving disease classification accuracy. Grad-CAMs that were employed illuminated the otherwise ‘black box’ of deep learning in medicine, allowing for increased physician confidence in adopting these new technologies in clinical practice.

**What is known:**

- Striking histopathological overlap exists between distinct but related conditions which poses a significant disease diagnostic challenge; such as for small bowel enteropathies including Environmental Enteropathy (EE) and Celiac Disease (CD).
- There is a major clinical need to develop computational [including Artificial Intelligence (AI) and deep learning] methods enabling clinicians to translate heterogeneous biomedical images into accurate and quantitative diagnostics.
- A major issue plaguing the use of AI in medicine is the so-called ‘black box’ of deep learning, an analogy which describes the lack of insight that humans have into how the models arrive at their decision-making

**What is new:**

- AI-based image analysis platform demonstrated high classification accuracy for small bowel enteropathies (EE vs. CD vs. histologically normal controls).
- Gradient-weighted Class Activation Mappings (Grad-CAMs) illuminated the otherwise ‘black box’ of deep learning in medicine, allowing for increased physician confidence in adopting these new technologies in clinical practice.

## Introduction

A major challenge of interpreting clinical biopsy images to diagnose disease is the often striking overlap in histopathology between distinct but related conditions.(1-3) Due to this significant clinical need, computational methods can pave the way for accurate and quantitative diagnostics(4). Computational modeling enhancements in medicine, particularly for image analysis, have shown the potential benefit of artificial intelligence (AI) for disease characterization.(5) There is an increasing interest in the utilization of deep learning,(6, 7) a subset of AI involving iterative optimization strategies based on pixel-by-pixel image evaluation.(8) These AI-based deep learning models, in particular Convolutional Neural Networks (CNNs), have been shown to be effective for medical image analysis.(5) CNNs have demonstrated potential for image feature extraction from diseases relying on radiological and histopathological diagnosis, particularly in the fields of ophthalmology and oncology.(9-13) Deep residual network (ResNet) is a CNN that has repeatedly shown success for image classification as it is optimized to gather fine grain attributes from regions of interest within the image.(14-16) It has also outperformed early deep learning models such as AlexNet(17) and VGG(18) and achieved superior performance on the ImageNet(19, 20) and COCO(21) image recognition benchmarks. While the deep learning models are noteworthy of their own accord, a major issue plaguing the use of AI in medicine is the so-called ‘black box’ of deep learning, an analogy which describes the lack of insight that humans have into how the models arrive at their decision-making.(22)

An accurate AI-based biopsy image analysis platform may enable efficient detection and differentiation of small bowel enteropathy damaged tissue architecture features which is challenging due to histopathological overlap.(2, 23-25) This will not only enable pathologists to filter and pre-populate scans, improving turn-around time, but also provide insight into previously uncharacterized tissue features unique to complex small bowel enteropathies such as Environmental Enteropathy (EE) and Celiac Disease (CD).

EE has been linked to poor sanitation and hygiene with dire consequences such as cognitive decline, oral vaccine response, and growth failure.(3) EE assessment and its histopathological differentiation from similar diseases (i.e. CD) is an essential task performed by pathologists.(26) Even though an EE scoring system has been proposed, it will benefit from approaches narrowing down specific cellular parameters important for an accurate EE diagnosis.(27) CD is an immune-mediation condition with sensitivity to gluten leading to small bowel injury. Modified Marsh score system(28, 29) has been used to classify the severity of CD but it does not account for complex disease features, such as the role of goblet or enteroendocrine cells, that can potentially improve diagnostic accuracy.

We have previously published a histopathological analysis model demonstrating 93.4% classification accuracy for identifying and differentiating between duodenal biopsies from children with EE and CD.(30) We also added a layer of explainability by using deconvolutions but they lacked biopsy regions of interest being specifically highlighted and did not fully explain the deep learning decision making process.(22) Despite increasing utilization of deep learning architectures in medicine, they remain underutilized for improving diagnostic accuracy and differentiation between histologically similar small bowel enteropathies. We now build upon our prior work to address knowledge gaps in the approaches previously reported. We aimed to: 1) optimize datasets by removing unavoidable bias acquired by archival data sourcing from multiple sites by using a hematoxylin and eosin (H&E) color normalization method with structure-preserving capabilities; 2) deploy an enteropathy focused deep learning CNN model with multiple layers modified to gather fine grain attributes from image regions of interest; 3) mimic the interpretation methodology of human pathologists by using a combination of multi-zoom and reduced parameter approaches (shallow deep learning models); and, 4) improve visualization of deep learning classification decision making.

## Methods

### Study Design and Archival Biopsy Image Dataset Acquisition

#### Archival Specimen Sources

Archival data for secondary analyses was sourced from: 1) primary Environmental Enteropathy (EE) studies from Pakistan (PK; Aga Khan University) and Zambia (ZA; University of Zambia School of Medicine, University Teaching Hospital), conducted 2013 to 2015,(31, 32) and 2) the University of Virginia (UVA), United States (US). UVA biopsies were from archival specimens with clinical diagnoses of CD and histologically normal controls (referred to as controls) from participants who had undergone esophagogastroduodenoscopy in the past 25 years (data accessed: 1992 to 2017) as part of an archival controls sub-study, methods reported elsewhere.(33) Biopsies available from each site for this project were archival and had been previously processed per local institutional H&E slide staining, tissue sectioning, and tissue paraffin embedding protocols. Our timeline for secondary data collection and analysis was from November 2017 to December 2019.

#### Biopsy Digitization

Biopsy slides for our secondary analysis had been previously digitized at high resolutions (average 20,000 by 20,000 pixels) to allow visualization of microscopic cellular features up to 40x (PK, US) and 20x (ZA) magnification. Digitization was done using Olympus VS 120 (Olympus Corporation Inc., Center Valley, Pennsylvania), Aperio Scanscope CS scanners (Leica Biosystems Division of Leica Microsystems Inc., Buffalo Grove, IL, United States), and Leica SCN400 brightfield scanner (Leica Microsystems CMS GmbH, Germany) at PK, ZA, and US, respectively. US archival biopsies were scanned using a prior detailed methods and work from these datasets has been published elsewhere.(2, 30, 32)

#### Archival Biopsy Image Data Acquisition

For the biopsy images acquired, diagnosis of EE was as previously defined in primary studies. At each site, clinical pathologists made diagnoses based on histological and clinical findings.(31, 32) For the US archival images, duodenal biopsy slides were obtained from the Biorepository and Tissue Research Facility at UVA with disease diagnoses as per clinical pathology reports. Biopsies from participants reported as controls were only included if there was no disease in any other part of the gastrointestinal tract (e.g., eosinophilic esophagitis, inflammatory bowel disease, *H. Pylori* gastritis, post-transplant liver disease etc.) or overall (e.g., patients with solid organ transplant, leukemia etc.).

### Biopsy Image Analysis Model Design

#### Dataset Pre-processing

##### Biopsy Image Patch Creation

High resolution whole slide images are patched for deep learning models to account for computational limitations.(34) Images were split into patches of 1000×1000 and 2000×2000 pixels with an overlap in horizontal and vertical axes of 750 and 1000 pixels, respectively. Patches that contained less than 50% tissue area were discarded and the rest were resized to 256×256 pixels (details noted in supplemental methods Figure S1). Each biopsy whole slide image generated an average of 250 1000×1000 and 40 2000×2000 patches.

##### Sample Size Augmentation, Balancing, and Justification

As there were more biopsy images for CD, EE and control images were up-sampled to balance datasets. Extensive data augmentations were performed for better generalization. As biopsy images exhibit both rotational and axial symmetry, a random combination of rotation (90, 180 or 270 degree angle), mirroring, and zoom (between 1x and 1·1x) was applied (Figure S2 in supplement). Deep learning studies have focused on obtaining as much image data as possible without any standard sample size recommendations.(34) There are limited studies focused on EE deep learning models and the sample size of this model is larger compared to our preliminary analysis.(30)

##### Stain Color Normalization

Stain color normalization using structure-preserving method as described by Vahadane et al.(35, 36) was used to eliminate bias due to color differences in the biopsies from different sites. It involved empirically selecting a target patch to normalize color across all patches (Figure 1). Three independent pathologists (LC, ZA, RI) completed a blind review of the color normalized biopsy images from different sites to assess the structure-preserving ability of the method.

**Figure 1:**
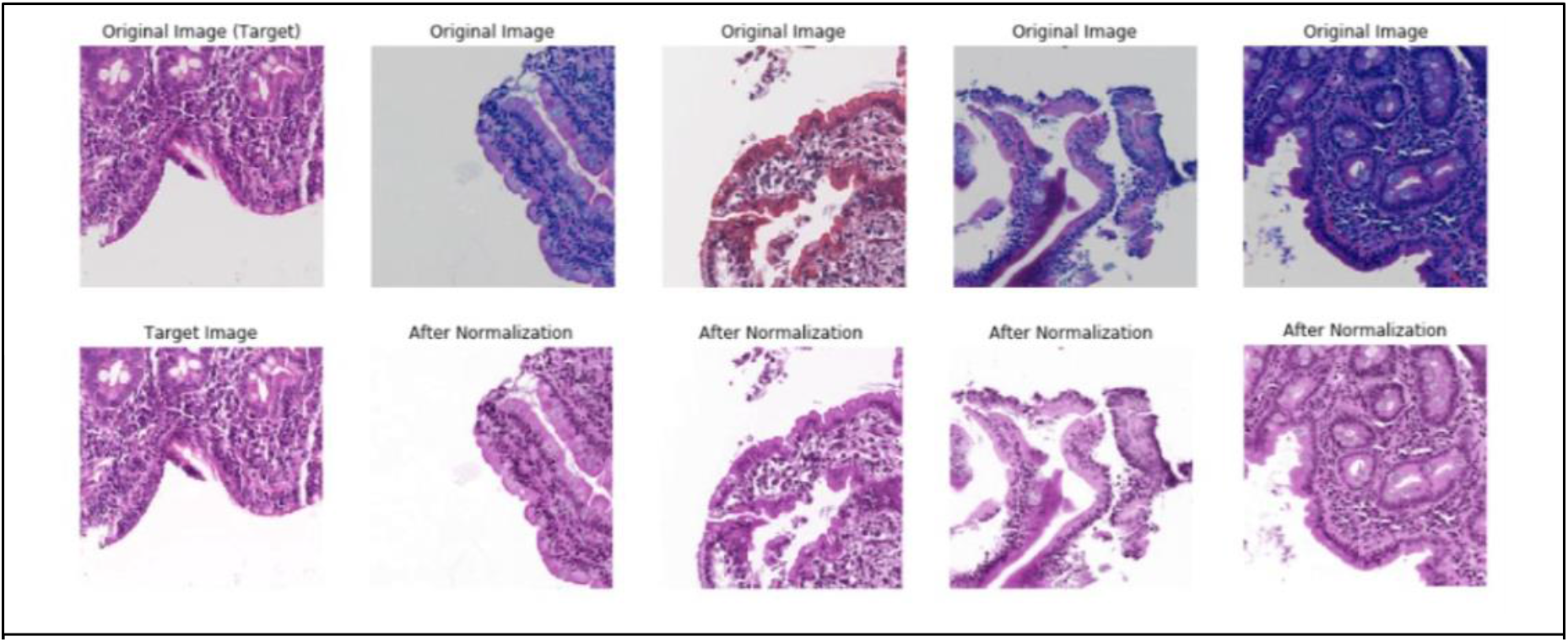
Stain Color Normalization - images in the top row highlight the differences in stain colors before normalization while the bottom row shows images post color normalization which was applied using the specified target image.

#### Image Analysis Model: Deep Learning Computer-Aided Biopsy Disease Classification System

Several deep learning Convolutional Neural Network (CNN) architectures were used to address specific questions as outlined below:

1. *Need for the identification of microscopic features visible at high magnification (zoom) level:* ResNet50 is a widely used deep CNN architecture with 50 layers for image classification requiring identification of microscopic patterns.(14-16) Final decision layers were modified to improve accuracy.(36) To combat data sparsity we used transfer learning, an established methods used to improve training using limited datasets by pre-training the model on the ImageNet dataset.(19, 37) Additional details of our modifications to the ResNet50 architecture are described in the Appendix S1 in supplement.
2. *Emulate human pathologist decision making process for visualizing biopsies at multiple levels of magnification (zoom):* we developed a framework incorporating multiple zoom levels (Figure 2). Biopsy images were segmented into 2000×2000 pixels then further into 1000×1000 pixel patches with an overlap of 750 pixels (horizontal and vertical axes). Two independent ResNet50 models were independently trained and the 1000×1000 and 2000×2000 corresponding patches were paired. Each pair was passed through the respective trained ResNet50 model and features from the last fully connected layer of each model were extracted and concatenated for an overall representation of specific regions of the biopsy images. This concatenated vector was further passed through a set of trainable linear layers for the final classification.
3. *Reduce computational model complexity used for disease classification:* Custom shallow CNN architecture was designed to help reduce the number of parameters the model optimized for disease classification. This Shallow CNN consisted of three convolutional layers (Figure 2; detailed methods in Appendix S2 as part of the Supplement).
4. *Explore methods of improving disease classification by using a combined model:* Ensemble models have been shown to generally improve the accuracy and robustness of classification.(38) We combined Resnet50 and shallow CNN to improve model classification accuracy. We obtained 3 values from the output layer of each CNN architecture which were then passed into a softmax function to obtain prediction probabilities for each patch being EE, CD or control. These predictions were then aggregated to acquire the final and better prediction for each biopsy image.

**Figure 2:**
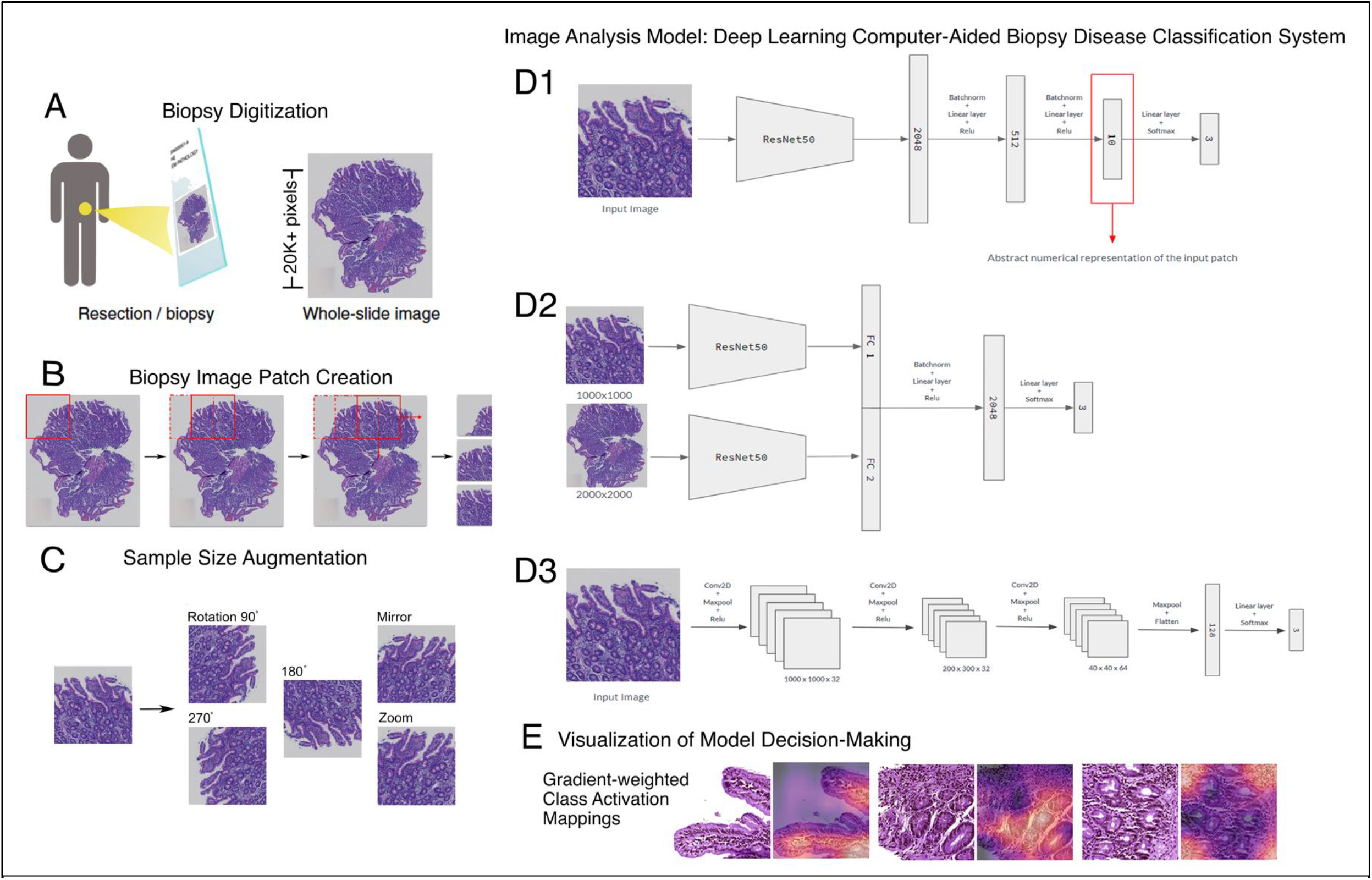
Combined figure explaining the methods for development of an image analysis model for diagnosis of small bowel enteropathies and black box feature detection: (A) Digitized duodenal biopsy slides were obtained for secondary analysis; (B and C) dataset was pre-processed by creation of biopsy image patches, sample size augmentation, and stain color normalization [shown in figure 1]; (D) development of the image analysis model with (D1) ResNet50, (D2) ResNet50 Multi-zoom Architecture, and (D3) custom Shallow Convolutional Neural Network (CNN); and, (E) visualization of model decision-making [explained in detail in Figure 4] Frameworks.

#### Visualization of Model Decision-Making

CNN activation maps were generated using Gradient-weighted Class Activation Mappings (Grad-CAMs)(39) to visualize the regions of interest utilized by the model for decision making (detailed methods outlined in Appendix S3 in the Supplement). These were reviewed by a team of medical professionals [pathologist specialized in gastroenterology (CM), pediatric gastroenterologist (SS)] enabling corroboration of the model results with incumbent classification. Since human intuition is required to assess if features highlighted can be biologically explained, we used the Environmental Enteropathic Dysfunction Biopsy lnitiative (EEDBI) scoring system for EE(27) and the modified Marsh score classification for CD(28, 29) to inform human medical professional intuition (Figure S3 and Table S1 in the Supplement).

#### Base Case for Comparison Using Existing Computer Vision Approach

An alternative method, CellProfiler,(40) using cellular feature extraction for explainability of the models was also explored (Appendix S4 and Figure S4 in the Supplement). CellProfiler isolated nucleated cells from the biopsy images and classified EE vs. CD vs. controls based on the cellular feature differences, achieving 65% accuracy.

### Ethical Considerations

The secondary analyses as part of this study were approved by University of Virginia Institutional Review Board. Ethical approval for prior original primary studies was obtained from: 1) the Ethical Review Committee of Aga Khan University, Karachi, PK (informed consent obtained from parents and/ or guardians for EE cases), 2) the Biomedical Research Ethics Committee of the University of Zambia School of Medicine, University Teaching Hospital, Lusaka, ZA (informed consent obtained from caregivers for EE cases), and 3) University of Virginia Institutional Review Board (waiver of consent granted).

This manuscript has been prepared in accordance with STARD guidelines.(41)

## Results

### Biopsy Image Dataset

We obtained 461 digitized biopsy images (171 H&E stained duodenal biopsy glass slides) from 150 participants (US: 124; PK: 10; ZA: 16). Our EE data consisted of 29 and 19 biopsy images from 10 PK and 16 ZA participants, respectively. Data from 124 UVA participants; 63 CD, 61 controls, was available for these analyses.

### Population Clinical Characteristics

Of the 150 participants, 77 (51·3%) were male. Median (inter-quartile range: IQR) age and LAZ/ HAZ (Length/ Height-for-Age Z score) of the EE participants was 22·2 (IQR: 20·8 to 23·4) months and -2·8 (IQR: -3·6 to -2·3), respectively (Table 1). Participants with EE were overall younger as compared to the US CD and controls (median age: 25·0; IQR: 16·5 to 41·0 months).

**Table 1:**
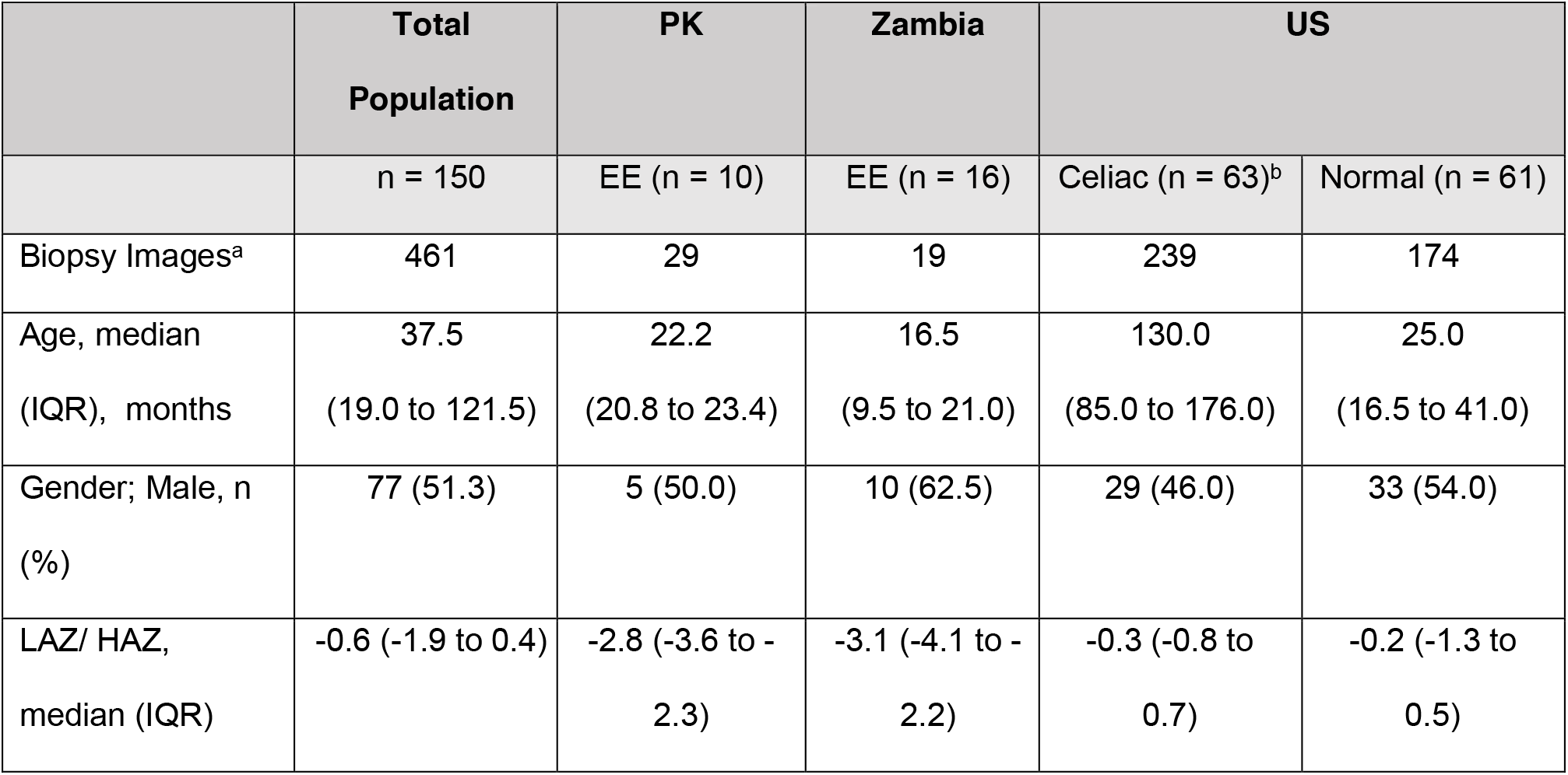
Population characteristics of the children from which biopsy images were obtained. Key: PK = Pakistan, Aga Khan University; Zambia = University of Zambia Medical Center, US = United States, University of Virginia; EE = Environmental Enteropathy; LAZ/HAZ = Length/Height-for-Age Z score; IQR = inter-quartile range (written as first quartile to third quartile) a – For some patients 2-3 biopsy images were available b – Height of 3 patients was not available for children with celiac disease (Z scores calculated for 60)

### Stain Color Normalization Assessment

Our panel of blinded independent pathologists confirmed that medically relevant cell types (lymphocytes, polymorphonuclear neutrophils, epithelial cells, eosinophils, goblet cells, paneth cells, and neuroendocrine cells) were preserved after color normalization. The panel also commented that features used for EE assessment via the published EE scoring system(27) were also visible. Although the granularity of eosinophilic cytoplasm and sharpness of paneth cell globules was less appreciated in post color-normalized images, these features were consistent throughout the biopsy images and were therefore hypothesized to not result in classification bias.

### Disease Classification Accuracy and Performance

Patches were used for training the models and the predictions were then aggregated to identify classifications for their parent biopsy images. Modified ResNet50 and Shallow CNN exhibited overall accuracies at the biopsy level of 98% (sensitivity: 93%, specificity: 94%) and 96% (sensitivity: 80%, specificity: 88%), respectively. The accuracy increased to 98·3% (sensitivity: 95%, specificity: 96%) with the ensemble. Accuracy of the multi-zoom ResNet50 architecture was 98% (sensitivity: 96%, specificity: 97%). Confusion matrices of the models’ classification for each disease are shown in Figure 4. Receiver operating curves (ROC) with area under the curve (AUC) were used to assess the certainty of the model classifications. Biopsy level AUC for modified ResNet50, ResNet50 with multi-zoom architecture, and shallow CNN were 0·99, 0·99, and 0·96, respectively (Figure 3). Error analysis and performance statistics for the models are included in Table S2 in the supplement.

**Figure 3:**
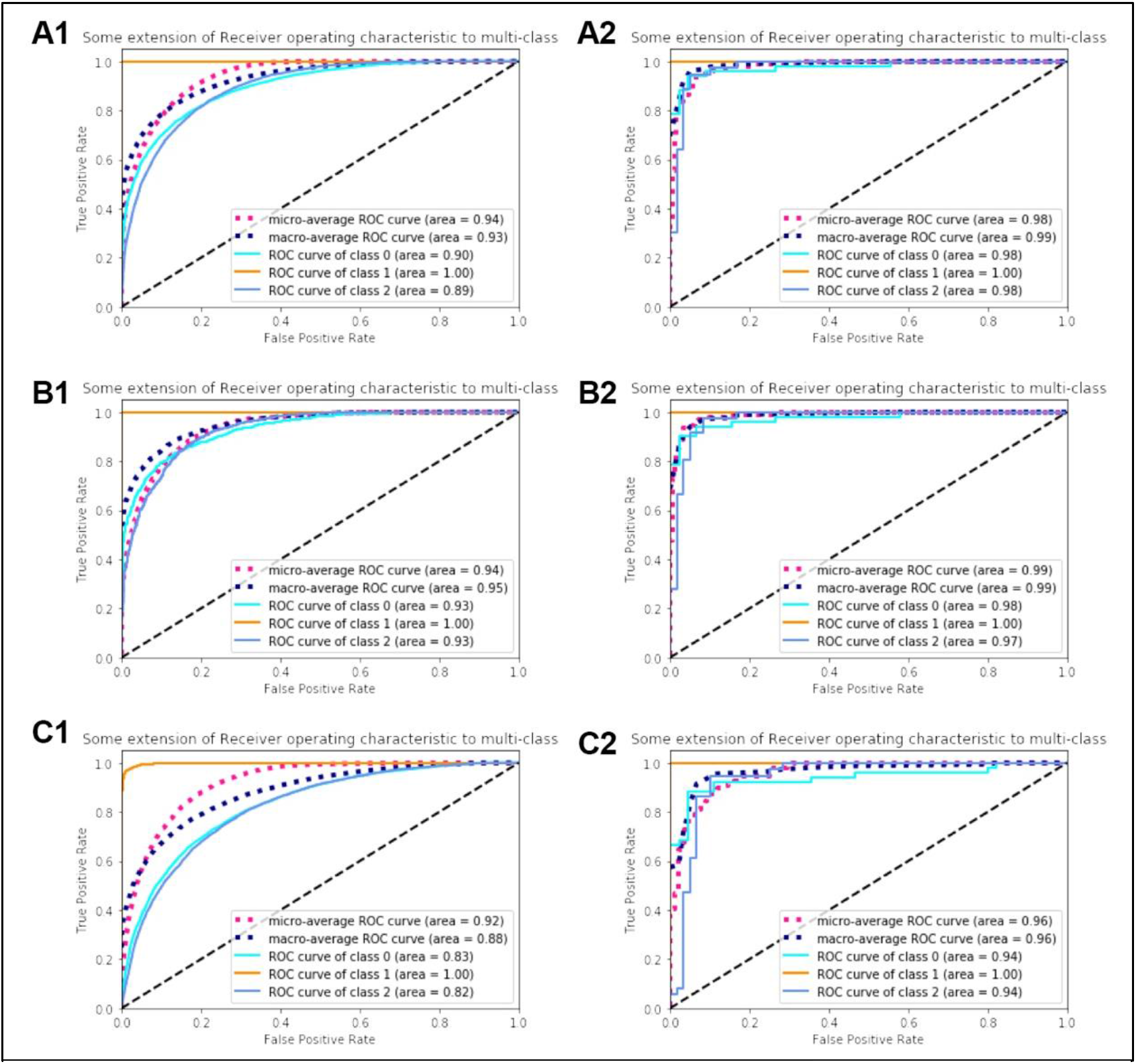
Accuracies of the deep learning models. A1, B1, and C1 show patch-level while A2, B2, and C2 show biopsy level accuracies of ResNet50, ResNet50 Multi-zoom Architecture, and Shallow CNN, respectively.

**Figure 4:**
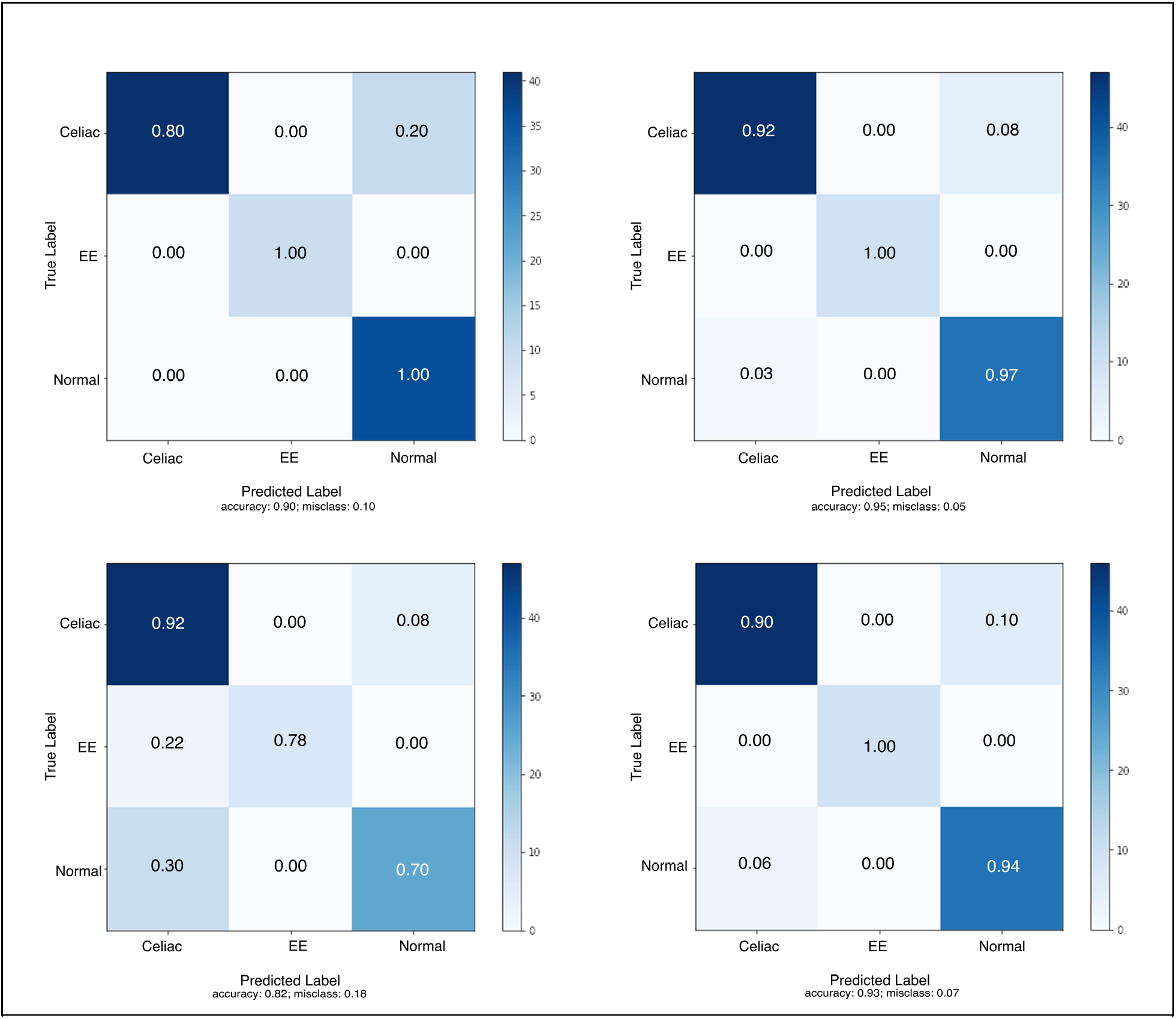
Confusion matrices for patch-level disease classification models (top left: ResNet50; top right: ResNet50 multi-zoom architecture; bottom left: Shallow Convolutional Neural Network (CNN); bottom right: ensemble. The numbers within the matrices represent normalized data in the form of percentages and darker colors indicate higher percentages.

### Grad-CAM Interpretations

Grad-CAMs were obtained for all the models (Figure 5). Modified ResNet50 and ResNet50 Multi-zoom mainly focused on similar areas for classification of EE vs CD vs controls while Shallow CNN focused on distinct, yet medically relevant cellular features (Table S3 in supplemental material for detailed Grad-CAM findings).

**Figure 5:**
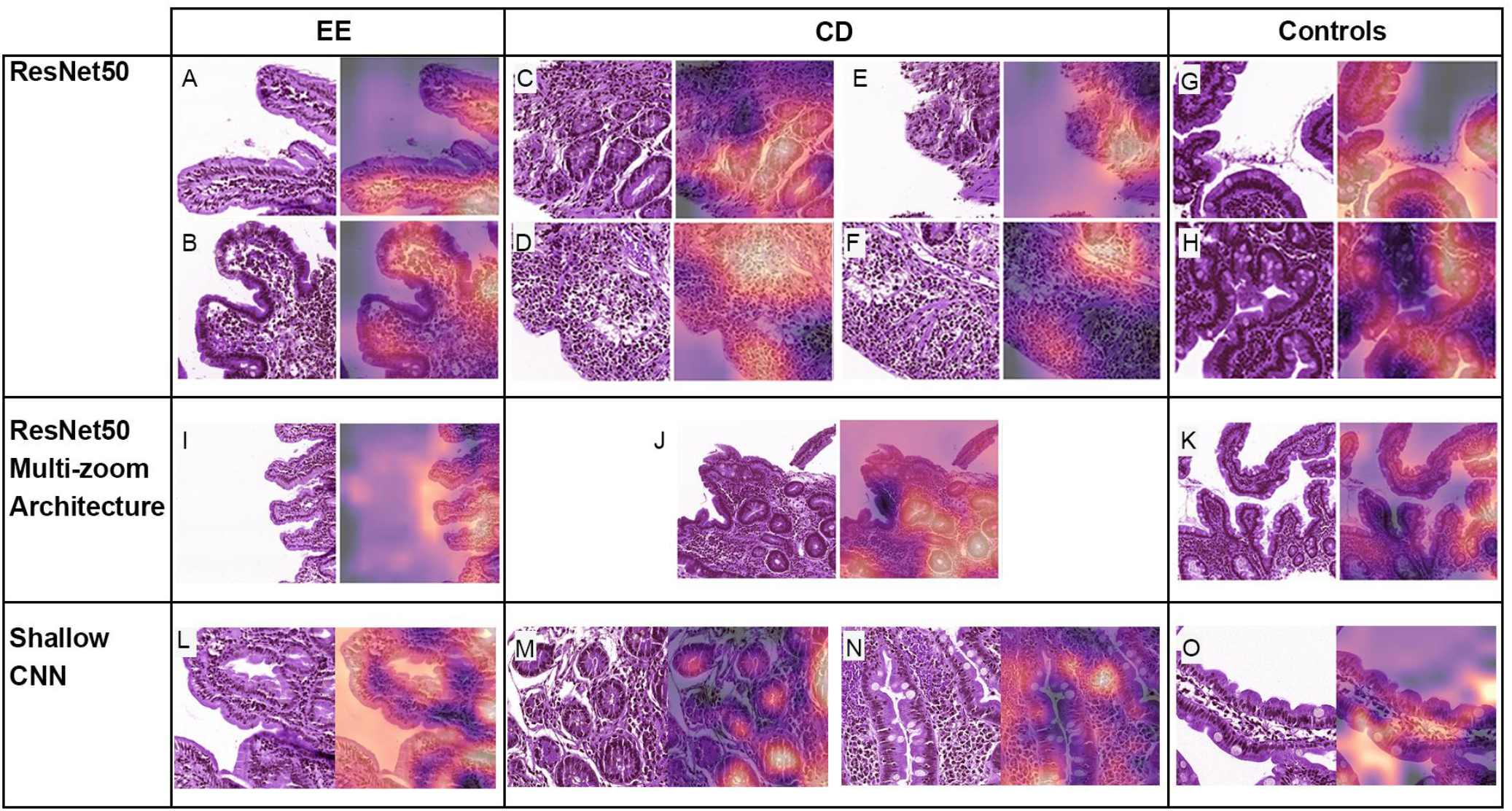
Gradient-weighted Class Activation Mappings for patches trained via modified ResNet50, ResNet50 Multi-zoom, and Shallow CNN are shown. Areas being highlighted are: A: superficial epithelium with high number of intraepithelial lymphocytes (IELs) and mononuclear cells in the lamina propria (LP); B: areas of white slit-like spaces representing artefactual separation of tissue within the LP and ignoring areas of crypt cross sections; C: crypt cross sections; D: areas with cells consistent with mononuclear cellular infiltrate; E: telescoping crypt; F: lymphocytes stacked in a row simulating a linear epithelial architecture; G: superficial epithelium with tall columnar cells and a brush border; H: base of crypts resembling superficial epithelium; I: superficial epithelium with high number of IELs; J: several crypt cross sections rather than 1 to 2 crypts when compared with ReNet50 only; K: superficial epithelium with abundant cytoplasm; L: surface epithelium with IELs and goblet cells; M: inner lumen of crypt cross sections; N: mononuclear cells within the LP; O: surface epithelium with epithelial cells containing abundant cytoplasm and goblet cells. Key: EE: Environmental Enteropathy; CD: Celiac Disease; CNN: Convolutional Neural Network

## Discussion

Diagnosis of overlapping histological diseases, such as small bowel enteropathies, lacks computational approaches for histopathological assessment.(30) While scoring systems such as the EE score and modified Marsh score for EE and CD, respectively, have the potential to diagnose and classify the severity of these diseases, there is a need to more accurately define the cellular parameters that play an important role in diagnosis.(27) We developed an AI-based image analysis platform, based on our preliminary results,(30) for the diagnosis of small bowel enteropathies. To the best of our knowledge, similar small bowel enteropathy image analysis platforms that include EE do not exist. We now build upon our prior work to address knowledge gaps in the approaches previously reported. Our modified ResNet50, ResNet50 with multi-zoom,

Shallow CNN, and an ensemble model (modified ResNet50 and Shallow CNN) depicted overall >90% classification accuracies. These AI-bases analytics and black box feature detection findings pave the way for standardizing and improving diagnoses for small bowel enteropathies.

Prior to training our deep learning models, dataset preparation included stain color normalization since with studies obtaining data from multiple sites for further analysis, an important consideration is to optimize datasets to remove site-specific bias.(35, 42) In our case, the biopsy images displayed visible stain color differences and the method we used to counter this bias has previously been proposed by Vahadane et al.(35) Along with stain color normalization, it successfully preserved small bowel architecture as assessed by our team of pathologists.

Our modified ResNet50 deep learning model trained to classify biopsy images of EE vs CD vs healthy controls was able to identify microscopic features of each disease. This work has been shown to be successful for other medical fields with ResNet providing superior performance for biopsy images.(14-16) This expands upon our preliminary model, as with the use of ResNet50 it provides powerful analytics for images. Further, the input to the previously published model(30) and other similar published studies for other clinical diseases consisted of small patches of whole slide images(15, 34) that represent approximately 2% of the original biopsy image. Such patch inputs do not take into account the fact that tissue features can be better visualized at multiple magnifications. The multi-zoom ResNet50 model employed in this study more closely emulates the process of human pathologists, who visualize biopsies at multiple magnification (zoom) levels for diagnoses.(43) The multi-zoom model’s Grad-CAMs focused on biologically similar areas as the model without multi-zoom, suggesting that the multi-zoom architecture does not bias the model significantly.

To further build upon our deep learning methods, we incorporated a shallow CNN and a final ensemble of ResNet and shallow CNN. The architecture of the shallow CNN was less complex than that of ResNet, a useful quality if an optimal computational environment is unavailable. A less complex model would also demonstrate utility if model parameters need to be reduced for a first-pass analysis of biopsy images to show proof of concept. Although the shallow CNN Grad-CAMs focused on far too many features for EE to be clinical useful, it focused on distinct and biologically informative features for CD. These Grad-CAM results led us to hypothesize that the shallow CNN would have better utility when combined with a more complex model such as ResNet50. This combination was manifested in the ensemble model, which showed an overall accuracy of 98.3%, higher than either model alone (ResNet50-only accuracy: 98%, Shallow-CNN-only accuracy: 96%).

While these results are noteworthy of their own accord, we further extracted Grad-CAMs to help illuminate the ‘black box’(22) of these deep learning models, to improve the insight into how the models arrive at their decision-making and provide the much needed ‘explainability’ for clinicians and researchers. Grad-CAMs supported the models’ use of biologically relevant features for defining each class: EE, CD or controls. The image features visualized via Grad-CAMs, if confirmed via molecular methods such as immunohistochemistry or RNA in situ hybridization, may pave the way for optimizing the EE and modified Marsh (for CD) scoring systems by drawing attention to the parameters pertinent to disease diagnoses.

Our secondary analysis built upon our earlier work by expanding and addressing the limitations. We computationally balanced the datasets and further augmented them to combat our prior issues with imbalanced datasets. In our preliminary work, we also utilized γ correction for stain color normalization without an assessment of the method’s structure-preserving ability. We have now utilized a more robust stain color normalization method that preserves structure, confirmed by a team of pathologists. We have also built upon the single model that we reported earlier to create a complex AI-based image analysis platform which utilizes multiple models to ensure a robust computational approach for histopathological image analysis. Lastly, our prior deconvolution-based approach to explaining the decision-making process of the preliminary model left much to be desired. Our current approach using Grad-CAMs to explain the models’ decision-making is a significant improvement, one more easily understood by the average medical professional.

Major strengths of our secondary analysis study include the addition of data from 48 new patients (in addition to the data reported for preliminary results) which increased the data volume for patch-level analysis. We also assessed the validity of our models with a wide variety of performance statistics (accuracy, sensitivity/ specificity, positive predictive value/ negative predictive value, precision, recall, and F1-score), many of which are often missing in the work done for deep learning in medicine. Our stain color normalization method for eliminating bias preserved tissue structure, an important quality for any successful normalization process. Our multi-zoom architecture emulating human pathologist biopsy assessment process, brings us one step closer to automated detection of biopsy disease features. Further, our use of Grad-CAMs for visualization of the models’ regions of interest will help both increase the confidence in AI-based decision making and also improve biopsy based small bowel enteropathy diagnosis.

Despite these strengths, we experienced several limitations. Since the data was obtained for secondary analysis, we were limited in terms of improving data quality due to differences in the scanners used for digitization and staining methods. Due to this we were unable to further modify the EE dataset to account for high classification accuracies. Despite this, we utilized an approach to potentially eliminate stain color differences. We were also limited in our ability to benchmark our findings due to limited literature for the use of deep learning models for small bowel architecture, especially the use of ResNet vs. custom deep learning approaches. To improve on our current efforts, more robust color normalization methods are underway. Finally, our techniques remain subject to the limitations inherent to tissue-based diagnostic and management approaches in clinical medicine, e.g., patchiness of findings and site specific workflows for tissue acquisition and processing.

## Conclusion

Artificial Intelligence-based analytics provide an exciting opportunity for improving small bowel enteropathy diagnostics with histological overlap. Our work suggests that models incorporated in our image analysis platform are capable of identifying biologically relevant microscopic features, emulating human pathologist decision making process, performing in the case of suboptimal computational environment, and being modified for improving disease classification accuracy. This work was improved by structure-preserving stain color normalization of the image inputs along with visualizations of the model outputs, which are imperative for determining the tissue features used for decision making. Further work will advance the clinical use of deep learning models for enteropathies and other histopathology based diseases, improving the effectiveness and efficiency of clinical care in gastroenterology and beyond.

## Data Availability

Data can be available on request

## Funding Information

Research reported in this manuscript was supported by National Institute of Diabetes and Digestive and Kidney Diseases of the National Institutes of Health under award number K23DK117061-01A1 (Syed), Bill and Melinda Gates Foundation under award numbers OPP1066203 (Ali) and OPP1066118 (Kelly), University of Virginia Center for Engineering in Medicine Grant (Syed and Brown), and University of Virginia THRIV Scholar Career Development Award (Syed). The content is solely the responsibility of the authors and does not necessarily represent the official views of the funding agencies.

Abbreviations
EE: Environmental Enteropathy
CD: Celiac Disease
AI: Artificial Intelligence
CNN: Convolutional Neural Networks
Grad-CAMs: Gradient-weighted Class Activation Mappings
ResNet: Deep residual network
H&E: Hematoxylin and Eosin
PK: Pakistan
ZA: Zambia
UVA: University of Virginia US: United States
EEDBI: Environmental Enteropathic Dysfunction Biopsy Initiative
IQR: inter-quartile range
LAZ/HAZ: Length/ Height-for-Age Z score
ROC: Receiver Operating Curves
AUC: Area Under the Curve

## Author Contributions

Concept and design: Syed, Brown, Shrivastava, Sengupta, Kant, Kang, Kelly, Amadi, Ali, Moore.

Acquisition, analysis, or interpretation of data: Syed, Ehsan, Khan, Guleria, Shrivastava, Sengupta, Kowsari, Cheng, Moskaluk, Kelly, Ali, Moore, Brown.

Drafting of the manuscript: Syed, Ehsan, Shrivastava, Sengupta.

Critical revision of the manuscript for important intellectual content: Syed, Guleria, Khan, Iqbal, Moskaluk, Kelly, Ali, Moore, Brown.

Statistical analysis: Syed, Shrivastava, Sengupta, Kowsari, Sali, Ehsan, Brown.

Obtained funding: Syed, Amadi, Ali, Moore, Brown.

Administrative, technical, or material support: Syed, Ehsan, Sadiq, Iqbal, Kelly, Ali.

Supervision: Syed, Kelly, Ali, Moore, Brown.

## Additional Contributions

We acknowledge the following individuals from Aga Khan University, Karachi, Pakistan, for contributing to this manuscript: Drs. Romana Idrees, MBBS, FCPS, and Zubair Ahmad, MBBS, FCPS, FRCPath, for assessment of stain color normalized biopsy images, Saad Mallick, medical student, for organization of image activation mappings, Dr. Saman Siddiqui, MD, for coordinating with the pathologists at Aga Khan University, and field workers (community health workers, led by Sadaf Jakhro, MSc, and coordinated by Dr. Tauseef Akhund, MBBS), data management unit (Najeeb Rahman, BS) and laboratory staff (Aneeta Hotwani, BS) for contributing to the data collection in this work. Patcharin Pramoonjago, PhD, Biorepository and Tissue Research Facility, University of Virginia, Charlottesville, assisted in data collection for this work. All individuals were compensated for their time.

## References

1. Sullivan PB, Marsh MN, Mirakian R, et al. Chronic diarrhea and malnutrition–histology of the small intestinal lesion. Journal of pediatric gastroenterology and nutrition. 1991;12(2):195–203.

2. Syed S, Yeruva S, Herrmann J, et al. Environmental enteropathy in undernourished Pakistani children: clinical and histomorphometric analyses. The American journal of tropical medicine and hygiene. 2018;98(6):1577–84.

3. Syed S, Ali A, Duggan C. Environmental enteric dysfunction in children: a review. Journal of pediatric gastroenterology and nutrition. 2016;63(1):6.

4. Topol EJ. High-performance medicine: the convergence of human and artificial intelligence. Nature medicine. 2019;25(1):44.

5. Sharma G, Carter A. Artificial Intelligence and the Pathologist: Future Frenemies? Archives of pathology & laboratory medicine. 2017;141(5):622–3.

6. Patel V, Khan MN, Shrivastava A, et al. Artificial intelligence applied to gastrointestinal diagnostics: a review. Journal of pediatric gastroenterology and nutrition. 2020;70(1):4–11.

7. Yang YJ, Bang CS. Application of artificial intelligence in gastroenterology. World J Gastroenterol. 2019;25(14):1666.

8. LeCun Y, Bengio Y, Hinton G. Deep learning. nature. 2015;521(7553):436.

9. Medeiros FA, Jammal AA, Thompson AC. From Machine to Machine: An OCT-Trained Deep Learning Algorithm for Objective Quantification of Glaucomatous Damage in Fundus Photographs. Ophthalmology. 2019;126(4):513–21.

10. Li Z, He Y, Keel S, et al. Efficacy of a Deep Learning System for Detecting Glaucomatous Optic Neuropathy Based on Color Fundus Photographs. Ophthalmology. 2018;125(8):1199–206.

11. Zhang Z, Xie Y, Xing F, et al., editors. Mdnet: A semantically and visually interpretable medical image diagnosis network. Proceedings of the IEEE conference on computer vision and pattern recognition; 2017.

12. Coudray N, Ocampo PS, Sakellaropoulos T, et al. Classification and mutation prediction from non–small cell lung cancer histopathology images using deep learning. Nature medicine. 2018;24(10):1559.

13. Beck AH, Sangoi AR, Leung S, et al. Systematic analysis of breast cancer morphology uncovers stromal features associated with survival. Science translational medicine. 2011;3(108):108ra13–ra13.

14. Bejnordi BE, Veta M, Van Diest PJ, et al. Diagnostic assessment of deep learning algorithms for detection of lymph node metastases in women with breast cancer. Jama. 2017;318(22):2199–210.

15. Wei JW, Wei JW, Jackson CR, et al. Automated detection of celiac disease on duodenal biopsy slides: A deep learning approach. Journal of pathology informatics. 2019;10.

16. Jiang Y, Chen L, Zhang H, et al. Breast cancer histopathological image classification using convolutional neural networks with small SE-ResNet module. PloS one. 2019;14(3).

17. Krizhevsky A, Sutskever I, Hinton GE, editors. Imagenet classification with deep convolutional neural networks. Advances in neural information processing systems; 2012.

18. Simonyan K, Zisserman A. Very deep convolutional networks for large-scale image recognition. arXiv preprint arXiv:14091556. 2014.

19. Deng J, Dong W, Socher R, et al., editors. Imagenet: A large-scale hierarchical image database. 2009 IEEE conference on computer vision and pattern recognition; 2009: Ieee.

20. Russakovsky O, Deng J, Su H, et al. Imagenet large scale visual recognition challenge. International journal of computer vision. 2015;115(3):211–52.

21. Lin T-Y, Maire M, Belongie S, et al., editors. Microsoft coco: Common objects in context. European conference on computer vision; 2014: Springer.

22. Castelvecchi D. Can we open the black box of AI? Nature News. 2016;538(7623):20.

23. Ramakrishna BS, Venkataraman S, Mukhopadhya A. Tropical malabsorption. Postgrad Med J. 2006;82(974):779–87.

24. Sullivan PB, Marsh MN, Mirakian R, et al. Chronic diarrhea and malnutrition–histology of the small intestinal lesion. J Pediatr Gastroenterol Nutr. 1991;12(2):195–203.

25. Pai RK, editor A practical approach to small bowel biopsy interpretation: celiac disease and its mimics. Seminars in diagnostic pathology; 2014: Elsevier.

26. Liu T-C, VanBuskirk K, Ali SA, et al. A novel histological index for evaluation of environmental enteric dysfunction identifies geographic-specific features of enteropathy among children with suboptimal growth. PLOS Neglected Tropical Diseases. 2020;14(1):e0007975.

27. Liu T-C, VanBuskirk K, Ali SA, et al. A novel histological index for evaluation of environmental enteric dysfunction identifies geographic-specific features of enteropathy among children with suboptimal growth. PLoS neglected tropical diseases. 2020;14(1):e0007975.

28. Modified Marsh Classification of histologic findings in celiac disease (Oberhuber) Stanford Medicine [Available from: http://surgpathcriteria.stanford.edu/gi/celiac-disease/marsh.html.

29. Oberhuber G. Histopathology of celiac disease. Biomedicine & pharmacotherapy. 2000;54(7):368–72.

30. Syed S, AI-Boni M, Khan MN, et al. Assessment of Machine Learning Detection of Environmental Enteropathy and Celiac Disease in ChildrenAssessment of Machine Learning Detection of Environmental Enteropathy and Celiac Disease in ChildrenAssessment of Machine Learning Detection of Environmental Enteropathy and Celiac Disease in Children. JAMA Network Open. 2019;2(6):e195822-e.

31. Iqbal NT, Sadiq K, Syed S, et al. Promising biomarkers of environmental enteric dysfunction: a prospective cohort study in Pakistani children. Scientific reports. 2018;8(1):2966.

32. Amadi B, Besa E, Zyambo K, et al. Impaired barrier function and autoantibody generation in malnutrition enteropathy in Zambia. EBioMedicine. 2017;22:191–9.

33. Iqbal NT, Syed S, Sadiq K, et al. Study of Environmental Enteropathy and MaInutrition (SEEM) in Pakistan: protocols for biopsy based biomarker discovery and validation. BMC pediatrics. 2019;19(1):247.

34. Iizuka O, Kanavati F, Kato K, et al. Deep Learning Models for Histopathological Classification of Gastric and colonic epithelial tumours. Scientific Reports. 2020; 10(1):1–11.

35. Vahadane A, Peng T, Sethi A, et al. Structure-preserving color normalization and sparse stain separation for histological images. IEEE transactions on medical imaging. 2016;35(8):1962–71.

36. Shrivastava A, Kant K, Sengupta S, et al., editors. Deep Learning for Visual Recognition of Environmental Enteropathy and Celiac Disease. 2019 IEEE EMBS lnternational Conference on Biomedical & Health lnformatics (BHl); 2019: IEEE.

37. Pan SJ, Yang Q. A survey on transfer learning. IEEE Transactions on knowledge and data engineering. 2009;22(10):1345–59.

38. Liu B, Cui Q, Jiang T, et al. A combinational feature selection and ensemble neural network method for classification of gene expression data. BMC bioinformatics. 2004;5(1):136.

39. Selvaraju RR, Cogswell M, Das A, et al., editors. Grad-cam: Visual explanations from deep networks via gradient-based localization. Proceedings of the IEEE lnternational Conference on Computer Vision; 2017.

40. Yu K, Zhang C, Berry G, et al. Predicting non-small cell lung cancer prognosis by fully automated microscopic pathology image features. Nat Commun. 2016; 7: 12474. Epub 2016/08/17. https://doi.org/10.1038/ncomms12474 PMlD: 27527408.

41. STARD 2015: An Updated List of Essential Items for Reporting Diagnostic Accuracy Studies: Enhancing the QUAlity and Transparency Of health Research; [Available from: https://www.equator-network.org/reporting-guidelines/stard/.

42. Khan AM, Rajpoot N, Treanor D, et al. A nonlinear mapping approach to stain normalization in digital histopathology images using image-specific color deconvolution. IEEE Transactions on Biomedical Engineering. 2014;61(6):1729–38.

43. Brunyé TT, Mercan E, Weaver DL, et al. Accuracy is in the eyes of the pathologist: the visual interpretive process and diagnostic accuracy with digital whole slide images. Journal of biomedical informatics. 2017;66:171–9.

